# Large Language Models for Social Determinants of Health Information Extraction from Clinical Notes – A Generalizable Approach across Institutions

**DOI:** 10.1101/2024.05.21.24307726

**Authors:** Vipina K. Keloth, Salih Selek, Qingyu Chen, Christopher Gilman, Sunyang Fu, Yifang Dang, Xinghan Chen, Xinyue Hu, Yujia Zhou, Huan He, Jungwei W. Fan, Karen Wang, Cynthia Brandt, Cui Tao, Hongfang Liu, Hua Xu

## Abstract

The consistent and persuasive evidence illustrating the influence of social determinants on health has prompted a growing realization throughout the health care sector that enhancing health and health equity will likely depend, at least to some extent, on addressing detrimental social determinants. However, detailed social determinants of health (SDoH) information is often buried within clinical narrative text in electronic health records (EHRs), necessitating natural language processing (NLP) methods to automatically extract these details. Most current NLP efforts for SDoH extraction have been limited, investigating on limited types of SDoH elements, deriving data from a single institution, focusing on specific patient cohorts or note types, with reduced focus on generalizability. This study aims to address these issues by creating cross-institutional corpora spanning different note types and healthcare systems, and developing and evaluating the generalizability of classification models, including novel large language models (LLMs), for detecting SDoH factors from diverse types of notes from four institutions: Harris County Psychiatric Center, University of Texas Physician Practice, Beth Israel Deaconess Medical Center, and Mayo Clinic. Four corpora of deidentified clinical notes were annotated with 21 SDoH factors at two levels: level 1 with SDoH factor types only and level 2 with SDoH factors along with associated values. Three traditional classification algorithms (XGBoost, TextCNN, Sentence BERT) and an instruction tuned LLM-based approach (LLaMA) were developed to identify multiple SDoH factors. Substantial variation was noted in SDoH documentation practices and label distributions based on patient cohorts, note types, and hospitals. The LLM achieved top performance with micro-averaged F1 scores over 0.9 on level 1 annotated corpora and an F1 over 0.84 on level 2 annotated corpora. While models performed well when trained and tested on individual datasets, cross-dataset generalization highlighted remaining obstacles. To foster collaboration, access to partial annotated corpora and models trained by merging all annotated datasets will be made available on the PhysioNet repository.

## INTRODUCTION

In the United States, social factors, including education, race, and poverty contribute to over a third of total deaths in a year [1]. According to a study that surveyed the number of deaths attributable to social factors in the United States in the year 2000, 245, 000 deaths were attributed to low education, 176, 000 to experiences of racism, 162,000 to low social support, 133,000 to individual-level poverty, 119, 000 to income inequality, and 39,000 to area-level poverty [1]. It should be noted that such mortality estimates are comparable to that of leading disease-related mortality rates. A growing body of research has reported on the negative impact of social determinants of health (SDoH) factors (socioeconomic and environmental factors that describe the conditions in which people are born, live, work, and age) creating undesirable circumstances, such as health disparities and discrimination [2-4]. For example, the likelihood of premature death increases as income goes down [2], children born to parents who haven’t completed high school are more likely to live in environments that contain barriers to health [3], and low education levels are directly correlated with low income, higher likelihood of smoking, and shorter life expectancy [2]. We note that in the context of this study the term SDoH is used in a broad sense covering not only socio-economic factors, but also behavioral, environmental, and other closely linked factors.

The mounting body of evidence demonstrating the impact of social determinants on health has resulted in an increasing acknowledgment within the health care sector that achieving improved health and health equity is likely to be contingent, at least in part, on addressing unfavorable social determinants [5-8]. Over the last decade, electronic health record (EHR) systems have been increasingly implemented at US hospitals and have generated large amounts of longitudinal patient data. These massive EHR databases are becoming enabling resources for diverse types of clinical, genomic, and translational research, with successful demonstrations by large initiatives such as the Electronic Medical Records and Genomics (eMERGE) Network [9], the Patient-Centered Outcomes Research Institute (PCORI) [10], and the Observational Health Data Science and Informatics (OHDSI) consortium [11]. However, most current studies do not use the full spectrum of EHR data, and more efforts have been devoted to including unstructured textual data in EHRs for real world evidence generation [12]. While some SDoH factors (e.g., race, sex) are present as structured data in EHRs, the lack of specific EHR fields to record SDoH information and the lack of standards for collecting data related to SDoH are some of the major reasons for insufficient SDoH documentation. On the other hand, clinical narratives contain more detailed characterization of several SDoH factors (e.g., living status, financial instability, social support, etc.), beyond the structured representation. The unstructured text data often contains detailed information to describe how multiple socio-economic, behavioral factors, and situational variables interact with each other.

Recent studies have utilized clinical notes to extract SDoH factors using NLP techniques [13-16]. The approaches range from utilizing rule-based techniques to deep-learning techniques and more recently large language models (LLMs) [16 17]. The problem is often formulated as a classification [16 18 19], entity recognition [20 21] or event extraction problem [15]. Majority of the existing work is limited to a single dataset from a specific institution or domain. In addition, these studies are limited to a small number of SDoH factors. Most existing NLP systems focus on factors such as smoking, drugs, alcohol, and housing [22]. Several other SDoH factors that have a profound impact on both physical and mental health (e.g., physical and sexual abuse, adverse childhood experience, financial issues, and social support) are less explored. Additionally, many of these factors have a temporal component which is often under emphasized in existing studies. The n2c2/UW SDOH Challenge [23] on SDoH extraction focused on the status, extent, and temporality of the factors, but was limited to five factors including the commonly studied alcohol, drug, tobacco, in addition to employment, and living situation using the Social History Annotated Corpus (SHAC) corpus [15].

The inclusion of SDoH in clinical notes can vary depending on various factors, including the healthcare system, individual clinician practices, and the specific context of patient encounters. These factors have a profound influence on the heterogeneity in the distribution of SDoH factors in clinical notes. Focusing on a specific note type, single institution data, specific hospital setting or individual documenting practices and developing datasets and testing NLP performance under these conditions might not always translate to a real-world scenario especially if developing models that generalize well across different settings is of prime importance. Hence, to explore deeper the influence of these variations, we curated multiple datasets covering the above factors.

To explore the single institution bias we collected data from four different hospitals and different hospital settings (inpatient and outpatient) including the publicly available MIMIC-III database. While “social history” sections are rich in SDoH (and many studies have solely used this information), extracting information from these sections alone might not always be ideal as SDoH factors can be scattered under different section headers depending on the type of notes, note templates, and individual note-taking styles of the providers. Hence, our datasets include psychosocial assessment notes, chart notes, social work notes, and all clinical notes from a published cohort study [24]. This variety in note types also accounts for the variety in documentation practices as they are written by physicians, nurses, social workers, etc.

What SDoH factors are less documented? Does this vary across medical specialties? For example, there is a better probability of finding some of the less studied SDoH factors such as social support, adverse childhood, and physical abuse-related information in clinical notes of patients experiencing mental health issues. These factors might be less prevalent in clinical notes written by physicians from other specialties, say Cardiology. Given such a real-world scenario, we perform a cross-institutional study exploring the variations in the distribution of SDoH factors and conduct experiments towards developing generalizable models that can extract multiple SDoH factors from clinical notes. Apart from traditional machine learning models and deep learning models we also inspect the use of an open-source LLM - LLaMA [25] for SDoH extraction and conduct a thorough evaluation of the model performance under different settings.

In summary, in this paper, we 1) explored the heterogeneity in the distribution of SDoH factors with regards to the healthcare setting, individual documentation practices, types of clinical notes and medical specialty; 2) designed and developed annotation guidelines to label sentences with multiple SDoH factors; 3) created four annotated corpora of SDoH with data from four healthcare systems; and 4) experimented with different text classification models to detect SDoH factors, including a LLM and evaluated their performance and showed how the varied label distribution of the SDoH factors across datasets impact cross-domain transfer learning.

To further test generalizability and encourage research and collaboration in this domain, we trained and evaluated models combining all the four datasets. The corresponding model will be made available on the PhysioNet repository for the research community.

## RELATED WORK

In clinical practice, there has been a lack of systematic collection and analysis of SDoH defined in the EHRs [14 19 26]. Consequently, structured data has insufficient documentation on SDoH. However, clinicians frequently include information about SDoH in their clinical notes as part of standard care [14]. To bridge this gap, NLP techniques have emerged to automatically identify SDoH information from unstructured notes. The current landscape of NLP approaches for identifying SDoH factors has been extensively reviewed [22 27].

Notably, Patra et al. conducted a comprehensive review of 82 NLP methods aimed at identifying SDoH [22]. These methods span various approaches, from rule-based to deep learning-based methods, presenting the identification of SDoH as either a classification problem [16 18 19] or a named entity recognition (NER) problem [15 20 21]. For example, Stemerman et al. [18] designed a multi-label classifier to identify six SDoH categories within sentences extracted from clinical notes sourced from the University of North Carolina’s clinical data warehouse. Similarly, previous studies explored the use of deep neural network architectures such as CNN [28], LSTM [29], and BERT [30] to automatically classify eight SDoH categories [16], using clinical notes categorized under “Social Work” in the MIMIC-III database [31]. In contrast, NER-based approaches, utilizing tools like cTAKES [23], CNN, and BERT-based models [24], were developed to extract SDoH factors from clinical text. Recently, encoder-decoder language models such as fine-tuned Flan-T5 XL and Flan-T5 XXL was used to extract 6 SDoH factors from narrative text in EHRs [17].

While multiple NLP methods are available, there are four primary challenges for SDoH identification. Firstly, many studies have focused on a limited set of SDoH factors. The review [22] revealed that among the 82 methods examined, only three SDoH factors were commonly addressed: smoking status (27 methods), substance use (21), and homelessness (20). Other critical factors such as education, insurance status, and broader social issues are still in the developmental stage. Secondly, a considerable number (22 out of 82) of these approaches rely on rule-based methodologies, which are highly dependent on specific lexicons. The lack of a standardized documentation framework for SDoH means these lexicons might not be easily transferrable or adaptable across different healthcare systems, significantly limiting the methods’ effectiveness. Recent work in this direction is the Gravity Project [32], a collaborative effort aimed at developing consensus-based data standards for SDoH information. Thirdly, most methods were developed and evaluated within specific healthcare systems, such as a Veterans Health Administration center [33], a Medical Center [34], and a Trauma Center [35]. The lack of broader testing raises concerns about the generalizability and adaptability of these NLP methods to diverse healthcare settings.

Finally, the decision to formulate the SDoH extraction task as classification vs. NER requires careful consideration. We decided to use multilabel classification as this approach offers flexibility in capturing the presence of various social determinants without requiring precise identification of entity boundaries. Annotators can focus on understanding the overall meaning and context of the sentence to determine which social determinants are relevant. This approach can potentially speed up the annotation process compared to the detailed entity-level annotation required in NER. Furthermore, there are certain factors such as social support, isolation or adverse childhood experiences that may not lend themselves well to strict entity identification as these factors often need to be inferred from the context. By adopting a multilabel sentence classification approach, annotators have the flexibility to capture these nuanced factors that cannot be precisely pinpointed as individual entities.

## METHODS

### Selection of SDoH factors

We performed an extensive review of literature to identify prior work on social determinants including systematic and scoping reviews [22 36] with focus on SDoH. Several ontologies/terminologies that cover SDoH factors [37 38] (e.g., Ontology of Medically Related Social Entities (OMRSE) [39], Semantic Mining of Activity, Social, and Health data (SMASH) system ontology [40]) were also analyzed to identify major concepts besides standards such as SNOMED CT and LOINC. Additional information was obtained from multiple surveys on SDoH including the All of Us SDoH Survey [41], Million Veteran Program (MVP) Lifestyle Survey [42], 2020 AHIMA Social Determinants of Health (SDOH) Survey [43], and the Gravity project [32]. Following this process with a preliminary analysis of clinical notes, we narrowed down to 21 SDoH factors which were then utilized for annotating clinical notes.

### Annotation

After obtaining the Institutional Review Board (IRB) approval for the study the datasets were annotated. The annotation process consists of two levels. At the first level (SDoH factors only) a sentence is assigned one or more labels from the 21 SDoH factors. For example, the sentence “*She is single, lives with her parents, works 3 days per week*.” will be labeled with *‘marital status*’, *‘living status*’ and *‘employment status*’. The second level of annotation (SDoH factors along with their corresponding values/attributes) provides more granular information about these factors with respect to their subtypes, presence or absence, and temporality. The sentence mentioned above at level 2 will be labeled with *‘marital status - single*’, *‘living status – with family*’ and *‘employment status – employed, current*’. If a sentence does not correspond to any SDoH factor, it was labeled ‘non SDoH’. Detailed information about the 21 SDoH factors and their attributes along with examples can be found in the annotation guidelines (see Supplementary Table T1 and T2 and Annotation Guidelines).

Two annotators (XC and XH) carried out the annotation with disagreements resolved by (VK) in consultation with the physician (SS). The annotators went through multiple rounds of training starting with a detailed discussion of the annotation guidelines and calculating the inter-annotator agreement (Cohen’s Kappa) after each training round. Depending on the datasets (discussed below), each round of annotator training utilized 10 clinical notes or 50 sentences. Once a Kappa value greater than 0.7 was achieved the remaining notes/sentences were annotated individually by the annotators and any discrepancies were resolved as mentioned above. The inter-annotator agreement after the final round of training was in the range of 0.73 - 0.90 for the datasets (which is similar to the annotator agreement reported in other studies [16]).

## Datasets

The prevalence of duplicate information in EHRs due to copy-pasting, templating, and summarizing is a major barrier to finding relevant information from EHRs [44 45]. A recent study reported that the duplication increased from 33% in 2015 to 54.2% for notes written in 2020 [44]. The over representation of certain SDoH factors because of the duplication impairs the training and evaluation of machine learning and deep learning models [46 47]. Hence, we removed duplicate sentences within multiple notes corresponding to a single patient from all datasets. This enabled us to reduce class imbalance among the SDoH factors to some extent. Nevertheless, the distribution of factors still varies, and class imbalance exists. We also observed several clinical notes documenting variations of the text “Nothing new to report” which were also eliminated.

We developed four datasets from de-identified and de-duplicated clinical notes of patients diagnosed with different health conditions from different hospitals. This includes “psychosocial assessment” notes of patients experiencing mental health issues, “chart notes” of patients diagnosed with Alzheimer’s disease, “social work” notes in the MIMIC-III database [31], and clinical notes of patients with chronic pain. The clinical notes were sampled using different techniques based on the note type and depending on where a majority of the SDoH information was present. IRB approval was obtained for this study prior to data collection. The details of these datasets are described below and summarized in Table 1.

**Table 1:** Statistics of the corpora used in this study.

| Corpus name | Care organization | Disease domain | Clinical note type | Type of setting | No. of notes | No. of patients | Time frame |
| --- | --- | --- | --- | --- | --- | --- | --- |
| HCPC | UTHealth Harris County Psychiatric Center | Mental health | Psychosocial assessment notes | Inpatient | 2000 | 1529 | 2001- 2021 |
| UTP | UTHealth Physicians | Alzheimer’s disease and related dementias | Chart notes | Outpatient | 1000 | 698 | 2007-2018 |
| MIMIC-III | Beth Israel Deaconess Medical Center | Intensive care | Social work notes | Inpatient | 500 | 264 | 2001-2012 |
| Mayo | Mayo Clinic and the Olmsted | Chronic pain | All clinical notes | Inpatient and outpatient | 527 | 62 | 2005-2015 |
|  | Medical<br>Center |  |  |  |  |  |  |

### Psychosocial assessment notes from UTHealth Harris County Psychiatric Center (HCPC)

Psychosocial assessments inform a comprehensive understanding of the psychological, social, and cultural context of a person guiding the development of individual care plans. Harris County Psychiatric Center (HCPC) is one of the largest providers of *inpatient* psychiatric care in the USA. About 10,000 patients are admitted yearly, including adults, adolescents, and children. Commonly treated conditions are psychotic or mood disorders, patients with acute crisis, and signs of endangering themselves or others. The EHR goes back to 2001 and includes about 120,000 unique patients. We randomly selected 2000 assessment notes (corresponding to 1529 patients) and extracted the “Social history” sections from these notes which are rich in SDoH information. These notes were then annotated by two annotators based on the annotation guidelines.

### Chart notes of patients diagnosed with Alzheimer’s disease from UT Physicians (UTP)

Several studies have shown the association of SDoH factors such as education level, isolation, and loneliness with the onset of Alzheimer’s disease and related dementias (ADRD) in older adults [48-50]. For the second dataset we utilized chart notes of patients diagnosed with (ADRD) from UT Physicians. UTP provides *outpatient* care with multiple satellite clinics throughout the greater Houston Area. Unlike the psychosocial assessment notes from HCPC where the majority of the SDoH factors were described in the “Social history” section, social history in the chart notes from UT Physicians (UTP) recorded mostly information related to substance use. Other SDoH information was scattered under sections titled “History of present illness”, “General observation”, etc. Additionally, chart notes are comparatively longer and contain information regarding medications and different body systems which are irrelevant to this study. Hence to increase the annotation efficiency and decrease the annotation time we developed a list of keywords to filter notes rich in SDoH information. The list of keywords was collected during the initial literature review and expanded by combining keywords from our prior work developing SDoH ontology and those obtained while annotating the clinical notes from HCPC [16 37 38 51].

### Social Work notes from MIMIC-III

The Medical Information Mart for Intensive Care (MIMIC-III) database contains more than two million free text notes under different categories (e.g., nursing notes, discharge summaries) with information of patients who stayed in critical care units of the Beth Israel Deaconess Medical Center between 2001 and 2012. It includes 2670 “social work” notes documenting the patient’s social and family history and the interactions of the social workers with the patient and family members during their stay at the hospital. We randomly selected and annotated 500 such notes.

### Clinical notes of patients with chronic pain from Mayo Clinic

For our fourth dataset we utilized a subset of the corpus from a study [24] that characterized chronic pain episodes in adult patients receiving treatment at Mayo Clinic and the Olmsted Medical Center. A total of 527 notes corresponding to 62 patients (retrieved using our keyword search) were annotated. These notes were mostly semi-structured having a template structure (especially for substance use) and duplication of information.

A schematic representation of the workflow is illustrated in Fig. 1. For brevity we will hereafter refer to the four datasets by the abbreviations of the names of the hospitals/database from which the notes were extracted as HCPC, UTP, MIMIC-III, and Mayo. During the annotation process, it was observed that some of the factors among the 21 annotated had a notably low prevalence and that those factors varied based on the corpus. To address the issue of class imbalance and enhance model performance, it is common practice to merge classes with fewer samples into a single category. Han et al. [16] employed this approach by consolidating their initial set of 14 annotation categories for SDoH classification into eight categories, with the less frequent ones merged into an “other-social” category. However, in our study, we chose not to follow this practice and instead trained our models on all classes, including those with a smaller number of samples. By doing so, we aimed to provide a more realistic evaluation of model performance in real-world scenarios, considering the wide variation in the distribution of SDoH factors. Furthermore, maintaining and training models on all classes is crucial for our cross-dataset evaluation to ensure consistent and reliable assessment, as any differences in the total number and types of classes would otherwise impact the evaluation process.

**Figure 1:**
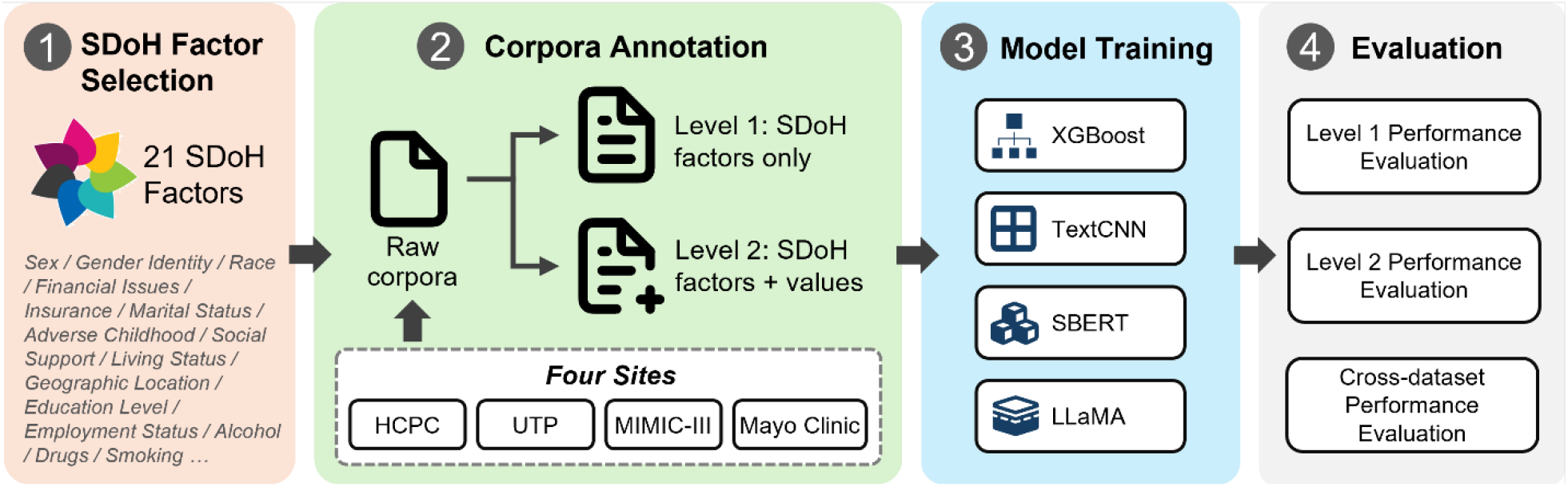
A schematic representation of the study workflow.

### Models

We experimented with four different models: XGBoost, TextCNN, SBERT and LLaMA. Since each sentence in the annotated datasets can potentially be annotated with multiple SDoH factors, we formulated the task of identifying SDoH factors in a sentence as a multi-label binary relevance problem for all models except LLaMA. Formally, given an input sentence *s* and a label set *C* of SDoH factors, a model produces a binary label of {*0, 1*} for each SDoH factor *c ∈ C*. We performed a supervised instruction finetuning for the LLaMA model. An instruction dataset was created with appropriate instructions to perform the task and the sentences and expected response as input and output to finetune the model. Below we provide a brief overview of these models.

#### XGBoost

We used the scikit-learn implementation of XGBoost [52] with tf-idf vectors as input features. OneVsRestClassifier was used with parameters n_jobs = -1 facilitating the use of all processors and max_depth value of 4.

#### TextCNN

We implemented a TextCNN model following [53] for multi-label text classification. We used pre-trained word embeddings (GloVe [54]) of dimension 300 for each word in a sentence as inputs. The model applies convolutional filters of kernel size 3, 4, and 5 to the input. The outputs of the convolutional filters are max-pooled and fed to a set of classification heads - each of them is a feed-forward network corresponding to a label.

#### SBERT

We used a pre-trained Sentence-BERT (SBERT) [55] encoder to encode each sentence into a dense vector representation aka. sentence embeddings. SBERT fine-tunes BERT [30] in a siamese/triplet network architecture that produces sentence embeddings that are semantically meaningful. We further fine-tuned SBERT for multi-label classification with the SDoH datasets. The output sentence embeddings of SBERT are passed to *C* binary classifier heads, each of which is a feed-forward neural network. Note that the SBERT sentence encoder is shared among all classifier heads. We fine-tuned the classifier heads and the shared SBERT encoder by minimizing the binary cross-entropy loss of all classifier heads. During inference, if a classifier head produces a score of 0.5 or more, the input sentence is assigned a positive label for that SDoH factor.

#### LlaMA

We utilized the open-source pretrained LLM, LLaMA 2 7B [22 56], and adapted it for multilabel classification by instruction finetuning. The training data for each dataset was converted into instruction demonstrations. An example of an instruction demonstration is shown in Supplementary Figures F1 and has three components – an instruction describing the task to perform, an input which in this case is the sentence from which SDoH factors needs to be extracted, and an output which is the gold annotation label(s) for that sentence. We utilized the approach mentioned in [57] using Hugging Face’s training framework with fully sharded data parallel and mixed precision training [58]. During inference, we provided the fine-tuned models with “instruction” and “input” to generate the “output”.

### Evaluation

In total we have eight annotated corpora – two each (level 1: SDoH factors only; level 2: SDoH factors and values/attributes) for each corpus. We evaluated the models on all corpora by first training and testing on the same corpus and then performing cross-dataset evaluation. To assess the model’s performance in handling the task of assigning multiple SDoH categories to a sentence, we computed micro-averaged precision (P), recall (R) and F1. Micro-average is calculated by considering the total number of true positives, false positives, and false negatives across all classes. **5-fold cross validation:** Each corpus was divided into 5 folds and performance of each fold was recorded and average performance across all 5 folds was calculated. Models were trained and tested only on the SDoH categories present in the corpus and hence the number of classes differs across the datasets.

#### Cross-dataset evaluation

It is observed that the performance is usually good for most models when trained and tested on the same corpus. We wanted to evaluate how different models would perform when trained on one corpus and evaluated on another. Each corpus was randomly divided into 7:1:2 ratio for train, validation, and test, respectively. Models were trained and tested on 22 classes (21 SDoH and 1 non SDoH) for level 1 annotated corpora and 71 classes (70 SDoH + values and 1 non SDoH) for level 2 annotated corpora.

#### Combined dataset evaluation

With factors having variations in their distribution across datasets, how would the performance be affected if we combine the datasets and train a single model? For evaluating this we retained the exact same split used for cross-dataset evaluation and combined the training splits of all four corpora at level 1 to create a single training split and combined the validation splits into a single validation split. The test splits were preserved separately. The same process was repeated for level 2 annotated corpora. Next, a model was trained on the combined training data, validated, and tested on the test splits separately for each corpus. The number of classes, similar to cross-dataset evaluation was 22 for level 1 and 71 for level 2.

## RESULTS

### Annotation and Datasets

#### HCPC

After eliminating duplicate notes/sentences, we ended up with a total of 4953 sentences (including 344 sentences with no SDoH). The distribution of level 1 annotated corpus is shown in Fig. 2 (top left). This dataset has a high prevalence of several factors such as adverse childhood, education level, isolation, geographic location where the person was born/raised, physical/sexual abuse and social support compared to other datasets in which these factors were not documented or less frequently documented. A total of 18 SDoH factors were annotated in this dataset.

**Figure 2:**
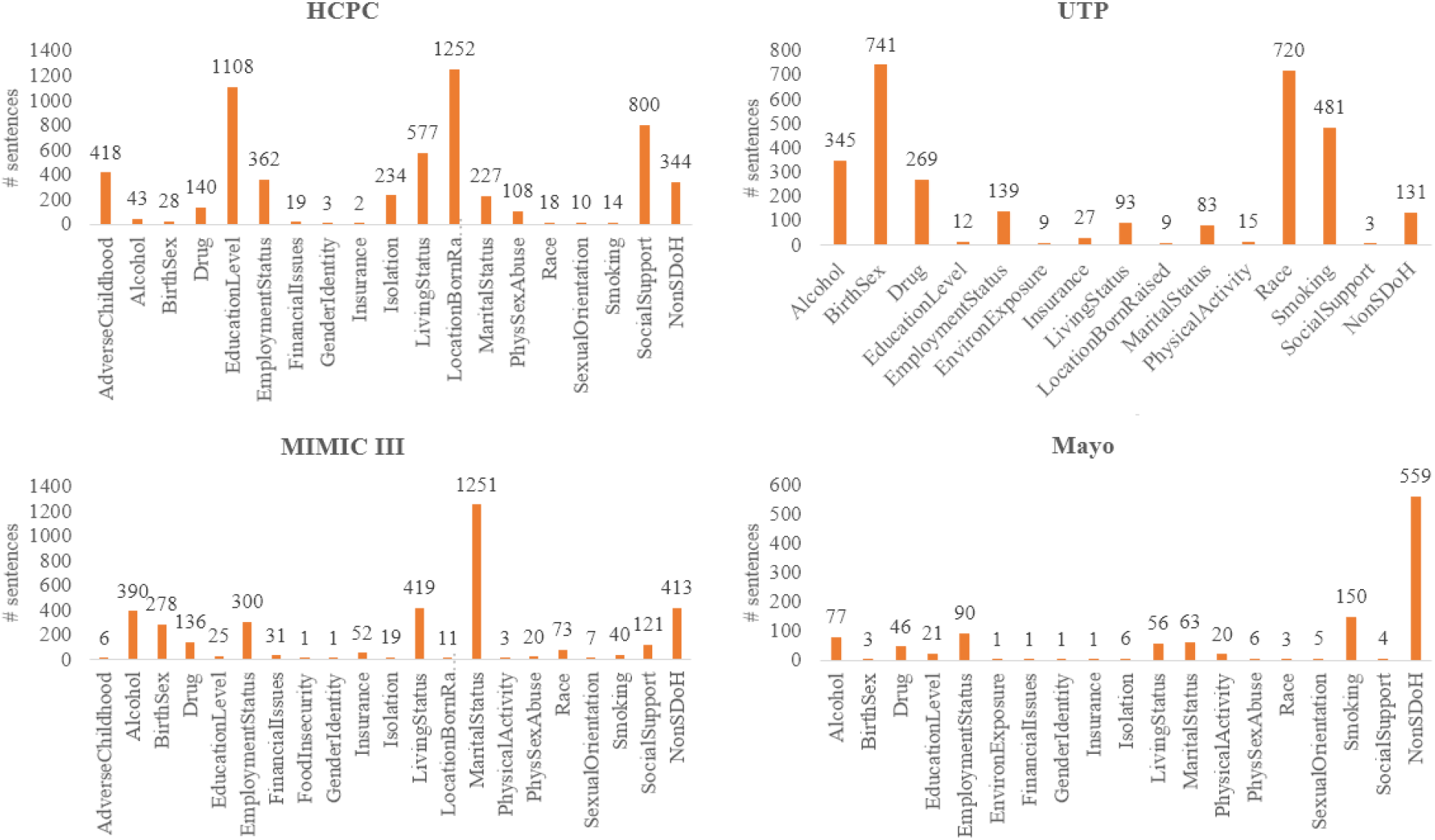
Distribution of SDoH factors for all four datasets (level 1 annotation).

#### UTP

A total of 1691 sentences were annotated from 1000 chart notes. Several of these notes had duplicate information because of copy-pasting for multiple visits by the same patient which were eliminated. For this dataset comprising of ADRD patients in an outpatient setting, the chart notes documented substance use information in detail under “social history” sections. Sex and race were almost always mentioned in the beginning (e.g., The patient is an 80 yr old Caucasian F …) followed by some information regarding the patient’s employment, living and marital status. 14 SDoH were annotated showing high prevalence of sex, race, and substance abuse.

#### MIMIC-III

As notes with note type “social work” were extracted from MIMIC-III database [31], these notes were written by social workers majorly documenting their interactions with patients’ families. This explains the towering frequency of marital status and some inference that could be made regarding living status and social support. Other factors that were frequently reported included alcohol and drug use, sex, and employment. A total of 2838 sentences were annotated from 500 notes with 20 SDoH factors present.

#### Mayo

This dataset was the smallest with 964 sentences annotated. The dataset contained all clinical notes of a chronic pain cohort with much of the data structured as templates to be filled in resulting in high volumes of duplicate information. In addition, major part was documentation of the diseases and medications explaining the high frequency of “non-SDoH” category. 18 SDoH factors were present with 10 categories having less than 10 samples.

The distribution SDoH factors with level 2 annotations for all the datasets are shown as part of Supplementary Figures F2-F5.

### Model performance

In this section, we show the results of our experiments with different models for all annotated corpora. Table 2 presents the results of 5-fold cross-validation on both level-1 and level-2 annotated corpora. Micro-averaged precision, recall and F1 scores are reported by averaging across all 5 folds. We observe that the instruction-tuned LLaMA model performed much better than all other models for all datasets. Compared to SBERT (second best performance), for level 1, LLaMA model had a performance increase in the range of 2.1% to 11.5%. The minimum increase was noted for UTP and the maximum for Mayo. For level 1, UTP dataset has only 15 classes (Fig. 2 top right) and has a comparatively better class balance. This is reflected in the model performance with 5-fold cross-validation consistently yielding an F1 score above 0.95 for all models. On the other hand, Mayo corpus (Fig. 2 bottom right) is highly imbalanced, is the smallest dataset, and has a total of 19 classes. It is interesting to note that the instruction tuned LLaMA model still achieved an F1 score of 0.94.

**Table 2.** Micro-averaged precision, recall, and F-1 metrics (average of 5-folds) of models across all datasets.

| Dataset | XGBoost | TextCNN | Sent. BERT | LLaMA |
| --- | --- | --- | --- | --- |
| **Level 1: SDoH factors only (P/R/F1)** |  |  |  |  |
| HCPC | 0.907/0.803/0.851 | 0.895/0.781/0.834 | 0.880/0.858/0.869 | 0.941/0.913/**0.927** |
| UTP | 0.982/0.935/0.958 | 0.980/0.927/0.952 | 0.979/0.948/0.963 | 0.990/0.979/**0.984** |
| MIMIC-III | 0.887/0.780/0.830 | 0.841/0.732/0.782 | 0.890/0.821/0.854 | 0.934/0.840/**0.883** |
| Mayo | 0.852/0.799/0.825 | 0.823/0.734/0.775 | 0.887/0.781/0.830 | 0.953/0.938/**0.945** |
| **Level 2: SDoH factors + values (P/R/F1)** |  |  |  |  |
| HCPC | 0.821/0.690/0.750 | 0.824/0.569/0.673 | 0.826/0.751/0.786 | 0.903/0.869/**0.886** |
| UTP | 0.946/0.880/0.912 | 0.889/0.815/0.850 | 0.957/0.882/0.918 | 0.982/0.932/**0.956** |
| MIMIC-III | 0.802/0.649/0.717 | 0.737/0.430/0.543 | 0.805/0.674/0.734 | 0.877/0.801/**0.837** |
| Mayo | 0.795/0.711/0.750 | 0.770/0.572/0.656 | 0.878/0.629/0.732 | 0.935/0.901/**0.918** |

If the number of classes increases and the number of datapoints remains the same, the model performance decreases as is evident from the results for level 2 annotated corpora (Table 2 bottom half). However, the decrease in performance for the LLaMA model (ranges from 2.7% to 4.6%) is less compared to other models (4.6 % to 11.3 % for XGBoost, 10.2% to 23.9% for TextCNN, and 4.5% to 12% for SBERT). For UTP, except for TextCNN all other models have an F1 score above

0.91. The maximum drop in performance for all models was noted when evaluated on the MIMIC-III corpus. This corpus is highly imbalanced (Supplementary Figure F4) with 1116 sentences having a label ‘*marital status – married*’.

### Generalizability Experiments

Figure 3 shows the performance of all models when trained on one corpus and evaluated on all other corpora (level 1 annotated corpora). The LLaMA model outperformed other models in cross-dataset generalizability. When finetuned on HCPC, LLaMA achieved an F1 score of 0.91 on both UTP and Mayo datasets and 0.82 on MIMIC-III, demonstrating an overall best performance on cross-dataset generalizability. Other notable performances include training on UTP and testing on Mayo (F1 = 0.82) and training on MIMIC-III and testing on UTP (F1 = 0.94). These results can be attributed to the high overlap of SDoH categories between these datasets, with a minimal impact from non-overlapping categories due to their limited sample sizes.

**Figure 3.**
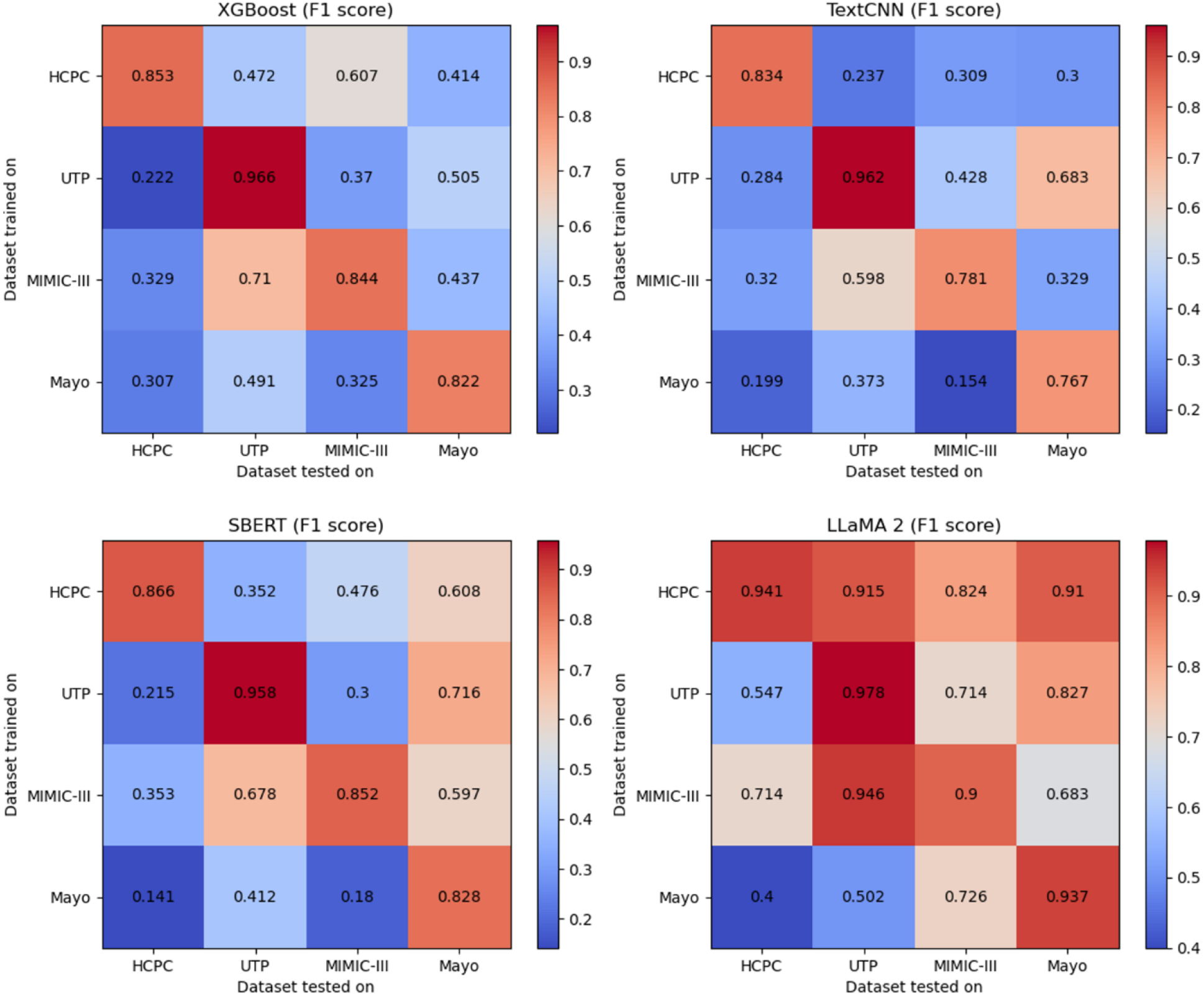
Heatmap of cross-dataset performance evaluation showing micro-averaged F1scores for all models on level 1 annotated corpora.

Even though with the increase in the number of classes the performance further decreased when evaluated for generalizability using level 2 annotated corpora, LLaMA model still performed better compared to other models (Fig. 4). Similar trends were observed as described with the level 1 annotated corpora. The lowest performance for any dataset pair was an F1score of 0.40 for LLaMA 2, 0.06 for SBERT, 0.04 for TextCNN, and 0.09 for XGBoost, clearly demonstrating the superior ability of instruction tuned LLaMA models to generalize better to unseen data. Precision and recall scores along with F1 for both levels of annotation is provided as Supplementary Tables T3 and T4.

**Figure 4.**
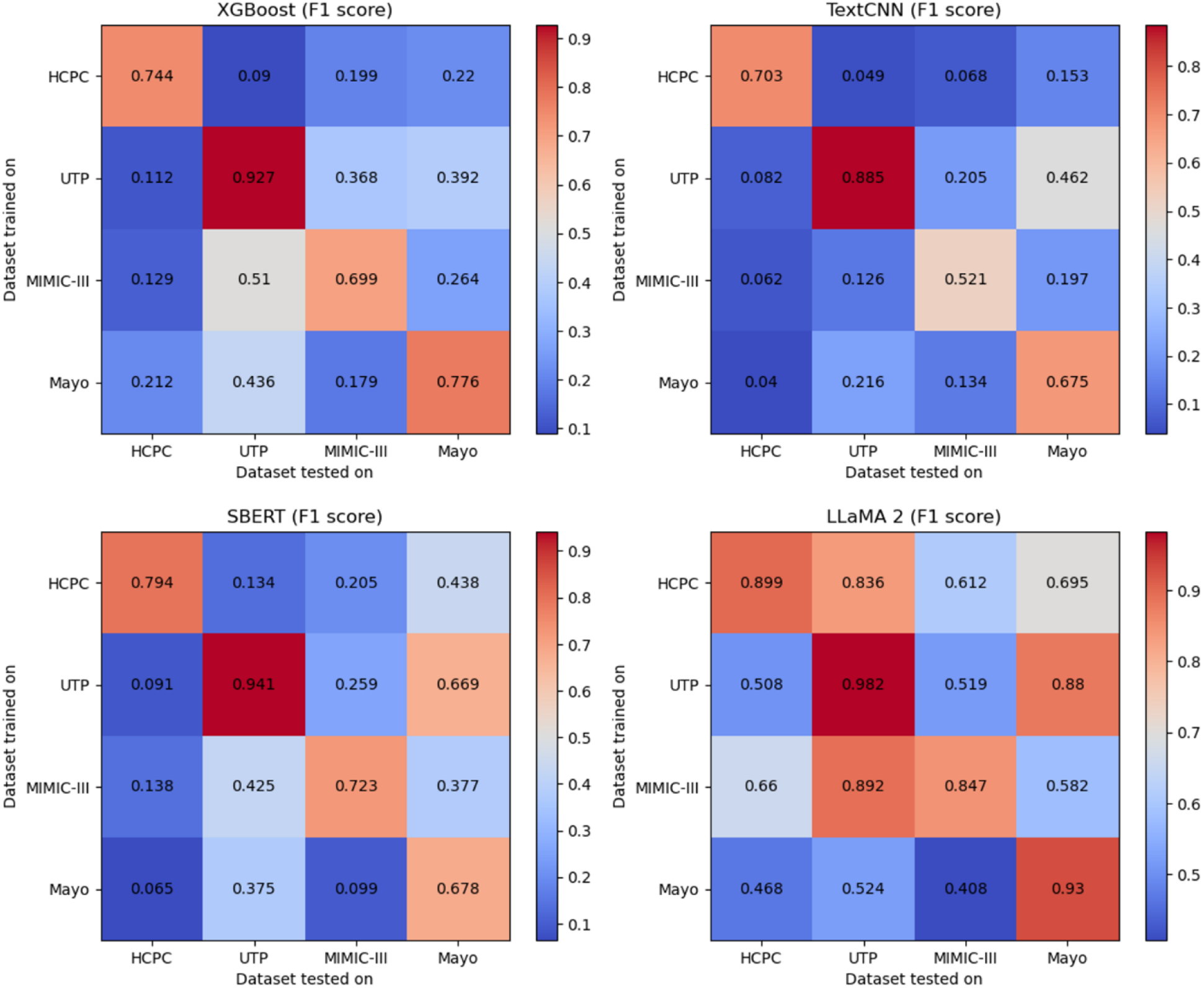
Heatmap of cross-dataset performance evaluation showing micro-averaged F1scores for all models on level 2 annotated corpora.

Final set of evaluations involved training models on a combined dataset and evaluating the performance of the resulting model on individual datasets. Almost similar performance is observed when comparing the results of combined training (Table 3) with that of training and testing on same dataset. The performance on Mayo dataset has improved for both XGBoost and SBERT models with more training data resulting in an increase of 5.5 % and 7%, respectively for level 1 annotation and 1.6% and 15.1% for level 2 annotation. On the contrary, LLaMA performance decreased considerably on the Mayo dataset.

**Table 3:**
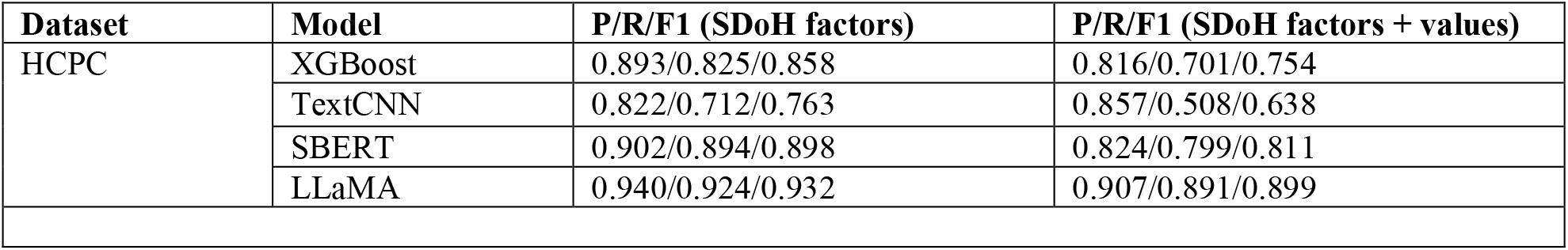
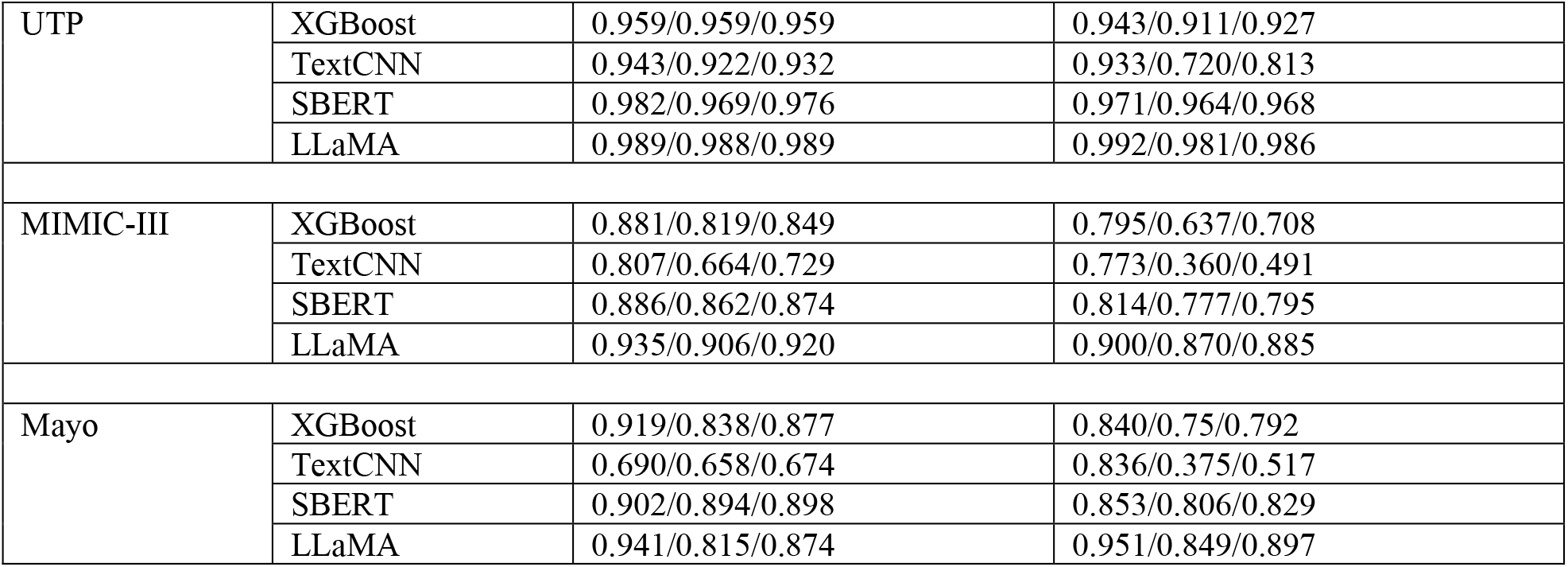
Performance of all models when trained on the combined dataset.

| Dataset | Model | P/R/F1 (SDoH factors) | P/R/F1 (SDoH factors + values) |
| --- | --- | --- | --- |
| HCPC | XGBoost | 0.893/0.825/0.858 | 0.816/0.701/0.754 |
|  | TextCNN | 0.822/0.712/0.763 | 0.857/0.508/0.638 |
|  | SBERT | 0.902/0.894/0.898 | 0.824/0.799/0.811 |
|  | LLaMA | 0.940/0.924/0.932 | 0.907/0.891/0.899 |
| UTP | XGBoost | 0.959/0.959/0.959 | 0.943/0.911/0.927 |
|  | TextCNN | 0.943/0.922/0.932 | 0.933/0.720/0.813 |
|  | SBERT | 0.982/0.969/0.976 | 0.971/0.964/0.968 |
|  | LLaMA | 0.989/0.988/0.989 | 0.992/0.981/0.986 |
| MIMIC-III | XGBoost | 0.881/0.819/0.849 | 0.795/0.637/0.708 |
|  | TextCNN | 0.807/0.664/0.729 | 0.773/0.360/0.491 |
|  | SBERT | 0.886/0.862/0.874 | 0.814/0.777/0.795 |
|  | LLaMA | 0.935/0.906/0.920 | 0.900/0.870/0.885 |
| Mayo | XGBoost | 0.919/0.838/0.877 | 0.840/0.75/0.792 |
|  | TextCNN | 0.690/0.658/0.674 | 0.836/0.375/0.517 |
|  | SBERT | 0.902/0.894/0.898 | 0.853/0.806/0.829 |
|  | LLaMA | 0.941/0.815/0.874 | 0.951/0.849/0.897 |

## DISCUSSION

In this paper, we undertook an extensive exploration of the heterogeneity in the distribution of SDoH factors within the healthcare setting. We investigated the interplay between various factors such as individual documentation practices, types of clinical notes, and medical specialty, aiming to understand the nuanced variations in the prevalence of SDoH factors across different contexts. To facilitate our investigation, we designed and developed annotation guidelines, which enabled us to label sentences containing multiple SDoH factors. Furthermore, we curated and constructed four annotated corpora of SDoH using data from four distinct healthcare systems, ensuring a diverse representation of SDoH factors. Leveraging these corpora, we conducted extensive experiments employing three text classification models and a generative model to detect SDoH factors. We not only evaluated the performance of these models but also highlighted the impact of the diverse label distribution of SDoH factors across datasets on the effectiveness of cross-domain transfer learning. We hope that our findings provide valuable insights into the complex landscape of SDoH factors, paving the way for enhanced understanding and future advancements in healthcare interventions and policies by facilitating and encouraging better documentation practices.

Our results demonstrate the ability of instruction tuned LLaMA model to consistently outperform all other models. The base LLaMA model being a general-domain LLM and the task of SDoH identification being a non-medical one might have rendered these models more suitable for this task compared to other biomedical NLP tasks which requires considerable biomedical knowledge. The performance of the XGBoost model demonstrates comparability to that of the SBERT model, and it is interesting to observe that the architectural variances did not significantly influence the outcomes when sufficient and diverse data is available, at least in this case within the scope of this study. The performance of TextCNN models was relatively much lower on two datasets - MIMIC-III and Mayo. MIMIC-III dataset has high lexical and semantic variability, and the sentences are diverse and do not follow any patterns/templates and is mostly written in a style of reproducing conversations with patients and their families, which might have affected the model performance. As for Mayo, it is the smallest dataset and has high class imbalance with more than half of the samples belonging to ‘*non SDoH*’ category.

LLMs require a substantial quantity of memory to store their parameters and intermediate computations. Training and inference with LLMs demand significant computational power that also translates to high energy consumption. While the parameter size and computational demands associated with LLMs are still challenging, these models consistently achieved greater than 0.9 micro-F1 on all datasets when finetuned and tested on the same dataset. But these models showed variations in performance and sometimes poor generalizability when tested across datasets. A thorough examination of the performance of the instruction-tuned LLaMA model and subsequent error analysis indicate the potential for further enhancement of the model’s performance. Analyzing the predictions, we found some cases where the model doesn’t make any predictions, makes predictions other than the expected class labels (e.g., 6^th^ grade instead of Education level) and not able to differentiate between SDoH pertaining to the patient from those referring to family members. The first two cases namely no responses and providing numbers such as date as response are the main reasons behind the considerably low performance on Mayo dataset when evaluated on the combined dataset (Table 3). Another interesting observation pertaining to the level of annotation was that if the sentence described the patient to be a non-smoker, then for level 1 annotated corpus, LLaMA did not generate ‘*smoking*’ as a label, whereas for the same sentence annotated at level 2 LLaMA consistently generated ‘*nonsmoker*’ as a response. These issues can be reduced to a certain extent by designing better prompts that has specific instructions to regulate such generations and providing detailed instructions from annotation guidelines as observed in our prior studies on LLMs [59]. In future, we plan to dive deep into these areas and explore their generative capabilities and potential to create synthetic datasets for SDoH covering a wide range of medical sub-domains for developing better generalizable models.

Keeping aside the performance of the LLaMA model and considering environments with resource limitations to train LLMs, among the other three models, even though SBERT and XGBoost models performed well when trained and tested on the same datasets, this performance did not translate when tested on other datasets. This low generalizability could be attributed to factors such as difference in 1) SDoH frequency distribution, and (2) the patterns in which SDoH factors are described differing significantly depending on the hospital setting, preference of individual practitioner, templates in place and medical specialties. To address this issue, data needs to be collected from diverse sources and combined to create datasets that capture the variations that contribute to heterogeneity. If the ultimate goal is to develop a single model (apart from an LLM) that can identify a variety of SDoH factors for a wide range of clinical notes originating from multiple institutions, federated learning [60] could be a promising direction. In federated learning, each institution will train their individual model with their privately-owned dataset and update their model through several rounds of model merging. After each round, the merged model will be broadcast to each institution before the next round of training begins. Other approaches such as a combination of Domain-incremental learning and Class-incremental learning [61] can also be applied to sequentially update a single model with new domains and new sets of classes. We will explore these in our future work.

With the increase in the number of classes resulting in high class imbalance there are not enough samples for the models to learn from, which is often the case with real-world data. It is to demonstrate the model performance in such cases, that we retained all the factors annotated without merging and tested the performance. It is impressive that LLaMA performed better than all models illustrating their potential to learn from fewer number of samples. Taking into consideration this situation and the generalizability issues observed, we developed and evaluated models by combining data from all four different settings. These models can be further finetuned with site specific data to suit the needs of specific institutions. We will make these models available on PhysioNet to encourage further research in this domain.

## CONCLUSION

Extracting and classifying social determinants of health from EHR and clinical notes have the potential for developing effective treatment plans, improving population health, and reducing health disparities. While what SDoH factors are documented and where they are documented varies widely across healthcare settings and medical specialties, the effects of these variations on the model performance in a real-world scenario is rarely studied, especially when developing generalizable models is a primary goal. Hence in this study, we attempt to provide an in-depth analysis into this heterogeneous distribution and analyze the performance of multiple models on SDoH extraction by designing and developing annotation guidelines for classifying SDoH factors and creating several annotated corpora using data from four healthcare systems. The datasets were curated to include different patient cohorts, note types, different layers of care in hospital settings, and documentation practices, thus showcasing the heterogeneity in the distribution of SDoH factors. Additionally, three classification models and a large language model were experimented with to detect SDoH factors with the LLM performing the best. The generalizability of the models across institutions was also evaluated. While all models perform relatively well when trained and tested on a single dataset, performance varied and dropped on cross-dataset evaluation, indicating the need for further research in this domain. To encourage research in this direction we will also make available models trained by combining all annotated datasets.

## Data Availability

All data produced in the present study are available upon reasonable request to the authors

## Acknowledgement

The authors thank Dr. Rajarshi Bhowmik for their suggestions and initial feedback in designing the study. This study was funded by the following grants - NIA 1RF1AG072799, R01AG084236, RM1HG011558 and R01AG083039. The funders played no role in study design, data collection, analysis and interpretation of data, or the writing of this manuscript.

## Data availability

The MIMIC III social work notes can be extracted after data user agreement from https://physionet.org/content/mimiciii/1.4/ and the MIMIC-III dataset used in the study will be made available on PhysioNet. The other datasets generated and/or analyzed during the current study are not publicly available due to privacy issues.

## Code availability

The instruction-tuned LLaMA models will be made available on PhysioNet website.

## Supplementary Materials

SDoH annotation guidelines, additional figures and tables are included along with the manuscript.

